# Knowledge and beliefs about dietary inorganic nitrate in a representative sample of adults from the United Kingdom

**DOI:** 10.1101/2024.07.24.24310912

**Authors:** Alex Griffiths, Evie Grainger, Jamie Matu, Shatha Alhulaefi, Eleanor Whyte, Eleanor Hayes, Kirsten Brandt, John C. Mathers, Mario Siervo, Oliver M. Shannon

**Author notes:** **Corresponding author:** Dr. Oliver Shannon. Human Nutrition & Exercise Research Centre, Centre for Healthier Lives, Population Health Sciences Institute, Newcastle University, Newcastle upon Tyne, NE2 4HH, UK. 01912081140. Shared first author (Alex Griffiths, Evie Grainger).

## Abstract

**Objective:** Evaluate knowledge and beliefs about dietary nitrate among United Kingdom (UK)-based adults.

**Design:** An online questionnaire was administered to evaluate knowledge/beliefs about dietary nitrate. Overall knowledge of dietary nitrate was quantified using a 21-point Nitrate Knowledge Index. Responses were compared between sociodemographic groups.

**Setting:** UK.

**Participants:** A nationally representative sample of three hundred adults.

**Results:** Only 19% of participants had heard of dietary nitrate prior to completing the questionnaire. Most participants (∼70%) were unsure about the effects of dietary nitrate on health parameters (e.g., blood pressure, cognitive function, cancer risk) or exercise performance. Most participants were unsure of the average population intake (78%) and acceptable daily intake (ADI) (83%) of nitrate. Knowledge of dietary sources of nitrate was generally low, with only ∼30% of participants correctly identifying foods with higher/lower nitrate contents. Almost none of the participants had deliberately purchased, or avoided purchasing, a food based around its nitrate content. Nitrate Knowledge Index scores were generally low (median[IQR]: 5[8]), but were significantly higher in individuals who were currently employed vs. unemployed (median[IQR]: 5[7]vs.4[7]; *p*<0.001), in those with previous nutritional education vs. no nutritional education (median[IQR]: 6[7]vs.4[8]; *p*=0.012), and in individuals who had heard of nitrate prior to completing the questionnaire vs. those who had not (median [IQR]: 9[8]vs.4 [7]; *p*<0.001).

**Conclusions:** This study demonstrates low knowledge around dietary nitrate in UK-based adults. Greater education around dietary nitrate may be valuable to help individuals make more informed decisions about their consumption of this compound.

## INTRODUCTION

Dietary inorganic nitrate is a water-soluble ion found in both plant- and animal-based foods ^(1,2)^. The prime exogenous source of dietary nitrate is vegetables, especially green leafy vegetables and beetroot. However, smaller amounts are also found in processed meat (in which nitrate is used as a preservative), certain fruits, legumes, herbs, and water ^(1,2)^. Additionally, in recent years, specifically formulated nitrate-containing supplements (e.g., nitrate-rich gels and concentrated beetroot juice ‘shots’) have become commercially available for individuals seeking to increase their intake of dietary nitrate above levels achieved through habitual diet alone ^(3,4)^.

Historically, dietary nitrate was considered to be an unwanted food contaminant with potentially deleterious health effects ^(5)^. In particular, higher intake of dietary nitrate was viewed as a risk factor for development of certain cancers and infant methemoglobinemia, which led to recommendations by public health bodies, including the World Health Organisation (WHO), to limit intake of this compound ^(6,7)^. However, with emerging evidence, a new perspective has begun to emerge which suggests that increased intake of nitrate may confer certain health benefits ^(4,8,9)^. Indeed, it has now been demonstrated that consumption of dietary nitrate in the form of vegetables/vegetable-derived products can improve markers of cardiovascular (e.g., reduced blood pressure and improved endothelial function) ^(10–14)^, brain (e.g., improved cognitive function and modulated cerebral blood flow) ^(15–19)^, and oral (e.g., modified oral microbiome and increased resilience against oral acidification) health ^(20–22)^. Similarly, nitrate has been shown to improve exercise capacity/performance across a range of population groups ^(23–25)^, making it a popular ergogenic aid amongst athletes ^(3,26)^.

Research interest in dietary nitrate has increased exponentially in recent years and in 2019 we demonstrated moderate knowledge of dietary nitrate amongst nutrition professionals, which was greatest in those possessing a PhD, indicating some dissemination of nitrate-based knowledge and contemporary research findings amongst nutrition professionals ^(27)^. Presently, there is no information around knowledge or beliefs about dietary nitrate in the general population. However, a better understanding of individuals knowledge/belief about dietary nitrate could be useful to: 1) help pinpoint specific areas where improved education or knowledge dissemination may be valuable (e.g., as part of public health campaigns aimed at promoting healthier eating); 2) identify areas where there may be misconceptions/misinformation about dietary nitrate, and help build strategies to counter these misconceptions/misinformation (e.g., by developing evidence-based resources, via science communication events and via engagement with traditional/social media); 3) recognise whether specific population subgroups differ in their knowledge/beliefs about nitrate, which could help identify target groups for educational resources; 4) help inform the design of behavioural interventions intended to modify dietary nitrate intake; 5) provide information which could be of value to manufacturers and retailers by helping understand whether knowledge/beliefs about dietary nitrate impact purchasing behaviour; and 6) serve as a reference point against which nitrate knowledge in the public could be tracked over time or compared against other population groups (e.g., different countries/athletes/clinical populations). Therefore, in this study we aimed to characterise knowledge and beliefs about dietary nitrate in a representative sample of UK adults. We also aimed to identify potential differences in knowledge/beliefs and between different population sub-groups. Our findings are likely to be relevant to researchers, policy makers, public health officials, and food manufacturers/retailers.

## METHODS

### Questionnaire Development and administration

We used a modified version of the KINDS questionnaire ^(27)^ to characterise knowledge and beliefs about dietary nitrate in the public. This questionnaire was previously developed by our group to evaluate knowledge/beliefs about dietary nitrate amongst nutrition professionals. Modifications were made to ensure appropriateness of language for a non-academic audience, removal of questions regarding biomarkers, metabolic processes and modification of guidelines limiting nitrate intake, and the inclusion of questions related to purchasing behaviour. The questionnaire was pilot tested with members of the public to ensure comprehensibility, and modifications were made to the wording and order of questions accordingly. The final questionnaire was sectioned into three parts: 1) Demographics (Table 1), 2) Knowledge and Beliefs (Table 2) and 3) Purchasing Behaviours (Table 2). Sub-categorising the questionnaire was advocated by participants involved in the pilot testing to improve readability and understanding. A final version of the questionnaire was built using an online survey tool (Online Surveys, Bristol, UK). The questionnaire was administered to a nationally representative sample of 300 participants (matched to the adult (>18 years) UK population regarding age, gender and ethnicity) in December 2022, via Prolific, an online crowd-sourcing platform that provides access to a pool of potential research participants (see^(28)^ for further details). Participants were given modest remuneration (£1.20) for their time, calculated to approximate a living wage (∼£10/h) on a pro-rata basis.

**Table 1.**
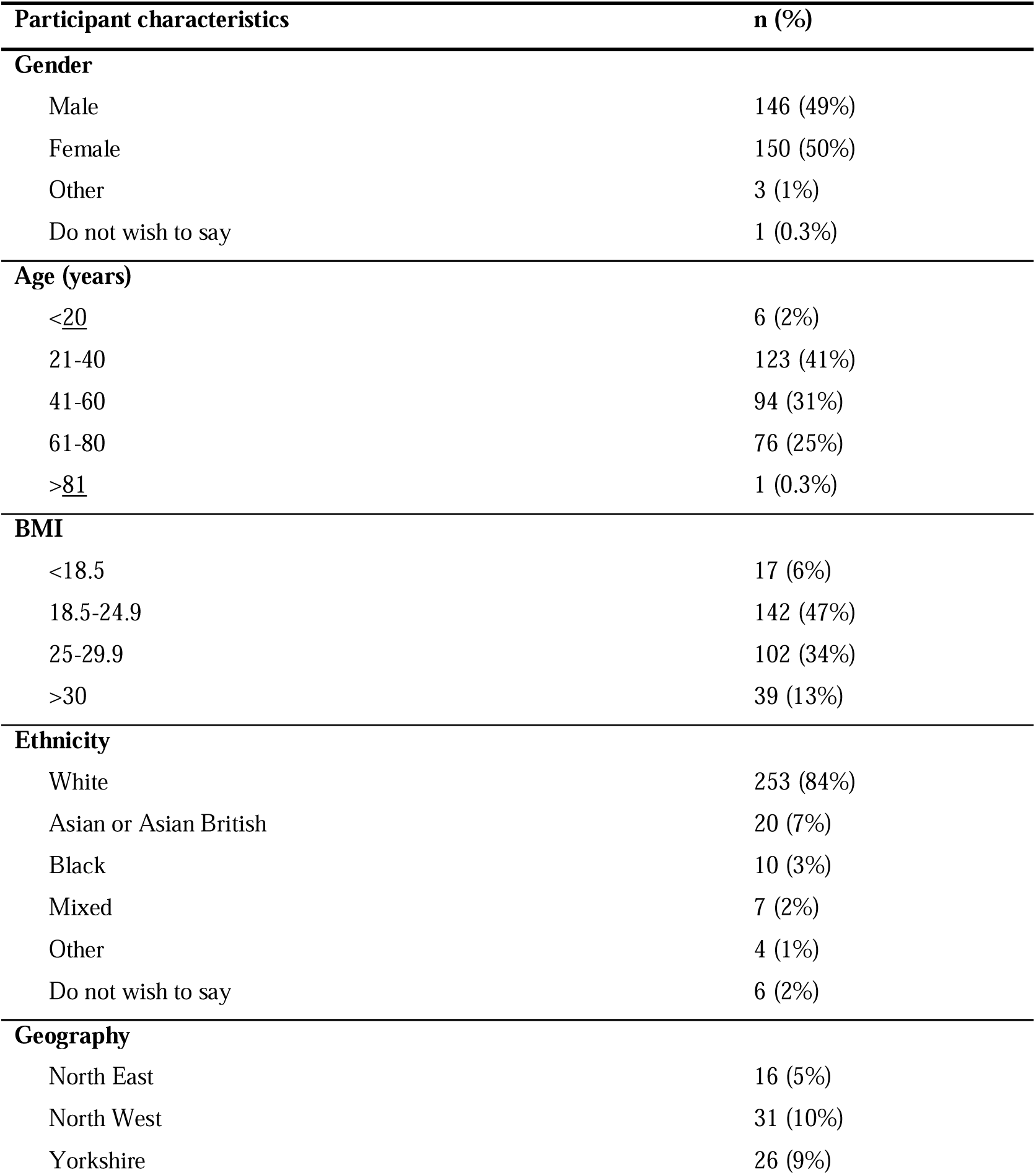
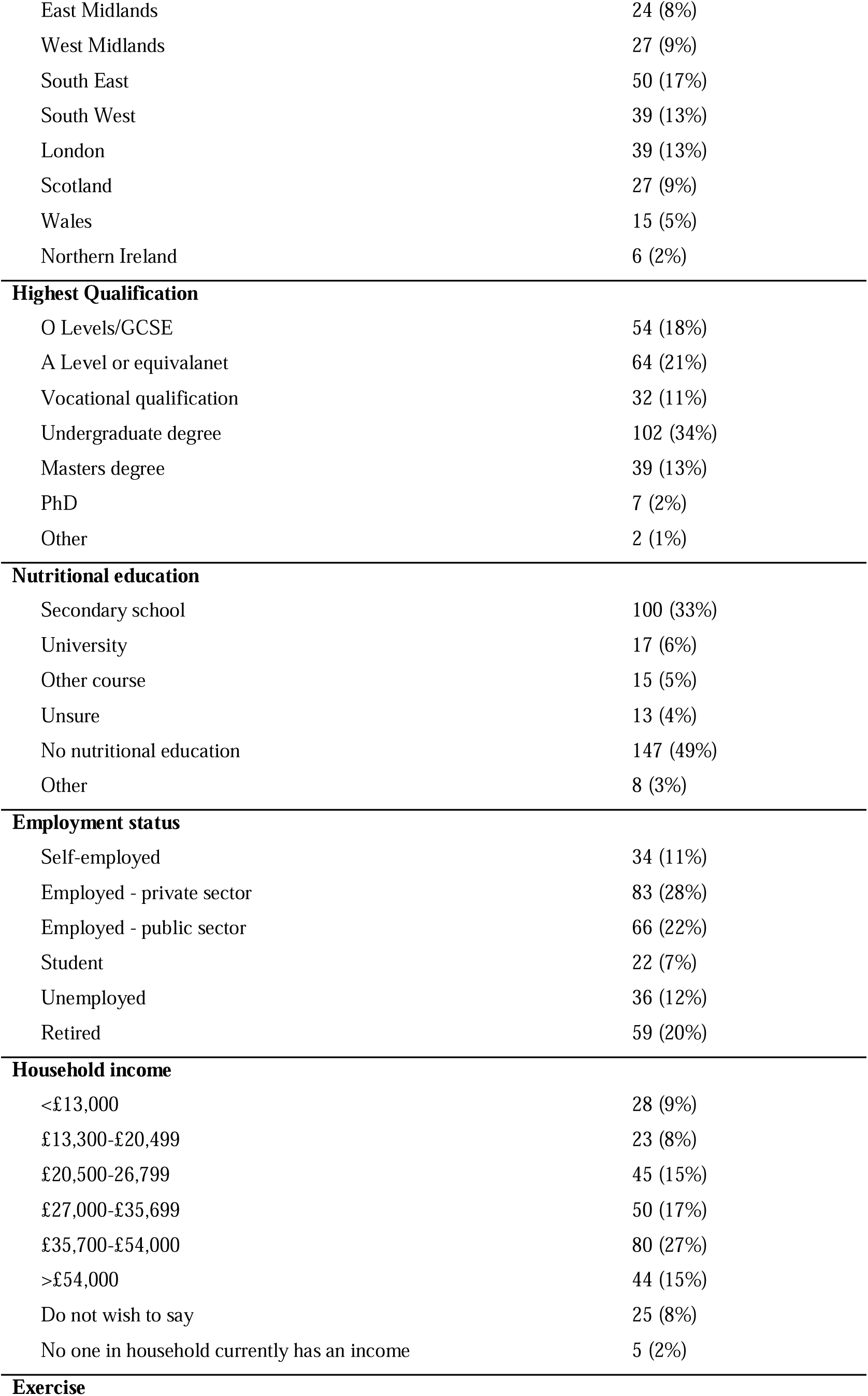
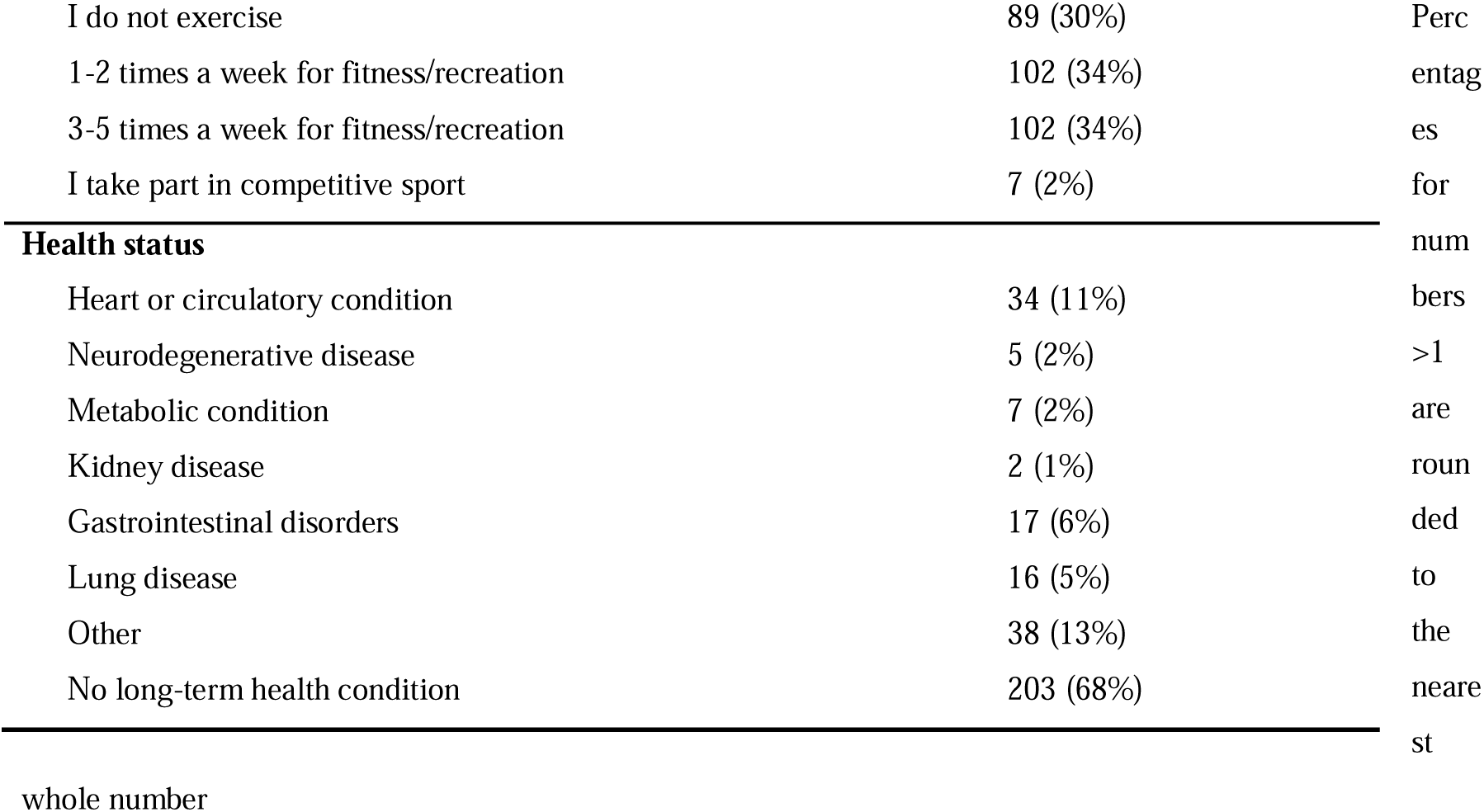
Participant characteristics.

| Participant characteristics | n (%) |
| --- | --- |
| **Gender** |  |
| Male | 146 (49%) |
| Female | 150 (50%) |
| Other | 3 (1%) |
| Do not wish to say | 1 (0.3%) |
| **Age (years)** |  |
| <20 | 6 (2%) |
| 21-40 | 123 (41%) |
| 41-60 | 94 (31%) |
| 61-80 | 76 (25%) |
| >81 | 1 (0.3%) |
| **BMI** |  |
| <18.5 | 17 (6%) |
| 18.5-24.9 | 142 (47%) |
| 25-29.9 | 102 (34%) |
| >30 | 39 (13%) |
| **Ethnicity** |  |
| White | 253 (84%) |
| Asian or Asian British | 20 (7%) |
| Black | 10 (3%) |
| Mixed | 7 (2%) |
| Other | 4 (1%) |
| Do not wish to say | 6 (2%) |
| **Geography** |  |
| North East | 16 (5%) |
| North West | 31 (10%) |
| Yorkshire | 26 (9%) |
| East Midlands | 24 (8%) |
| West Midlands | 27 (9%) |
| South East | 50 (17%) |
| South West | 39 (13%) |
| London | 39 (13%) |
| Scotland | 27 (9%) |
| Wales | 15 (5%) |
| Northern Ireland | 6 (2%) |
| **Highest Qualification** |  |
| O Levels/GCSE | 54 (18%) |
| A Level or equivalent | 64 (21%) |
| Vocational qualification | 32 (11%) |
| Undergraduate degree | 102 (34%) |
| Masters degree | 39 (13%) |
| PhD | 7 (2%) |
| Other | 2 (1%) |
| **Nutritional education** |  |
| Secondary school | 100 (33%) |
| University | 17 (6%) |
| Other course | 15 (5%) |
| Unsure | 13 (4%) |
| No nutritional education | 147 (49%) |
| Other | 8 (3%) |
| **Employment status** |  |
| Self-employed | 34 (11%) |
| Employed - private sector | 83 (28%) |
| Employed - public sector | 66 (22%) |
| Student | 22 (7%) |
| Unemployed | 36 (12%) |
| Retired | 59 (20%) |
| **Household income** |  |
| <£13,000 | 28 (9%) |
| £13,300-£20,499 | 23 (8%) |
| £20,500-26,799 | 45 (15%) |
| £27,000-£35,699 | 50 (17%) |
| £35,700-£54,000 | 80 (27%) |
| >£54,000 | 44 (15%) |
| Do not wish to say | 25 (8%) |
| No one in household currently has an income | 5 (2%) |
| **Exercise** |  |

|  |  |  |
| --- | --- | --- |
| I do not exercise | 89 (30%) | Perc |
| 1-2 times a week for fitness/recreation | 102 (34%) | entag |
| 3-5 times a week for fitness/recreation | 102 (34%) | es |
| I take part in competitive sport | 7 (2%) | for |
| **Health status** |  | num |
| Heart or circulatory condition | 34 (11%) | bers |
| Neurodegenerative disease | 5 (2%) | >1 |
| Metabolic condition | 7 (2%) | are |
| Kidney disease | 2 (1%) | roun |
| Gastrointestinal disorders | 17 (6%) | ded |
| Lung disease | 16 (5%) | to |
| Other | 38 (13%) | the |
| No long-term health condition | 203 (68%) | neare |
|  |  | st |

whole number

**Table 2.**
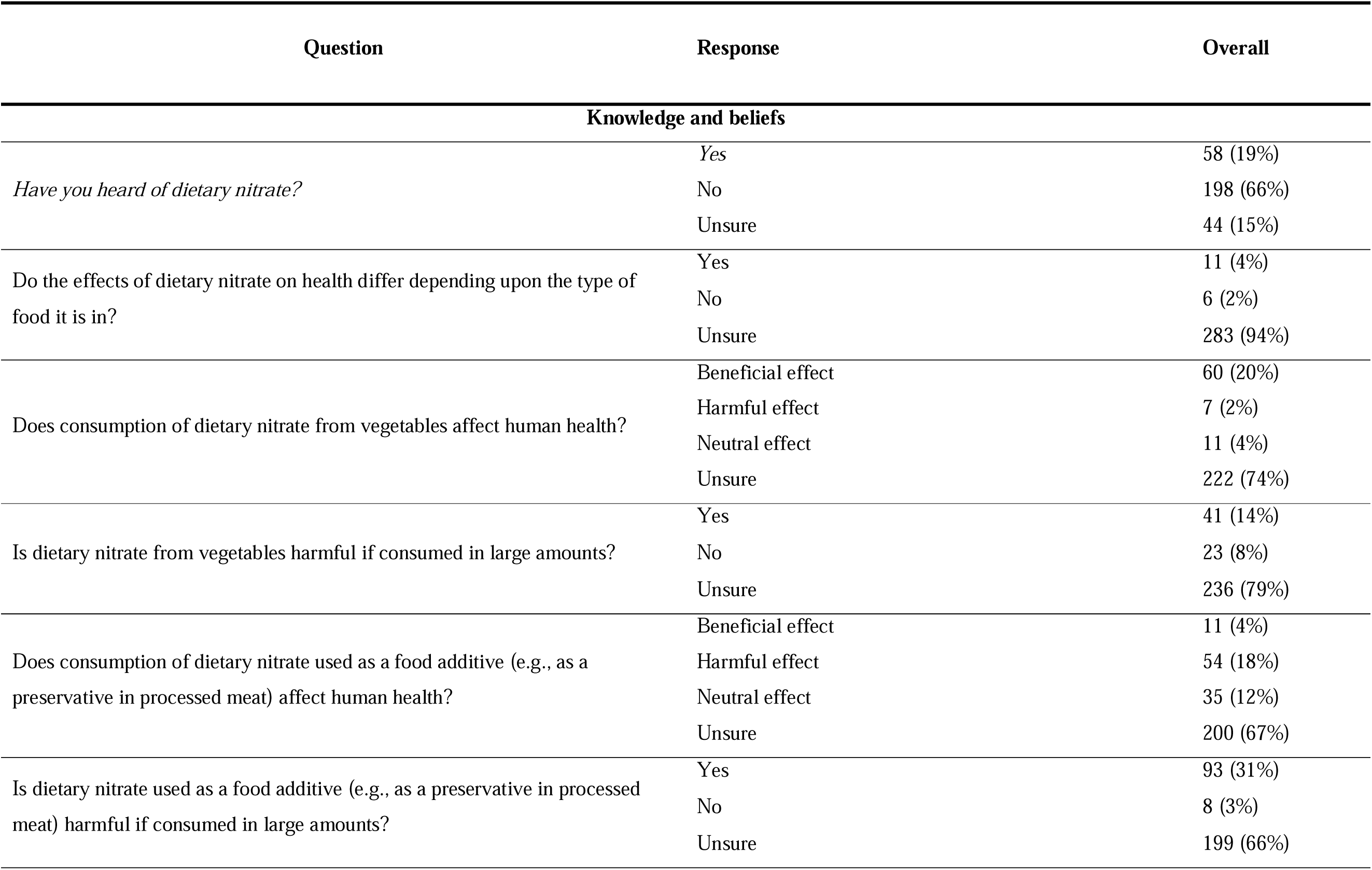
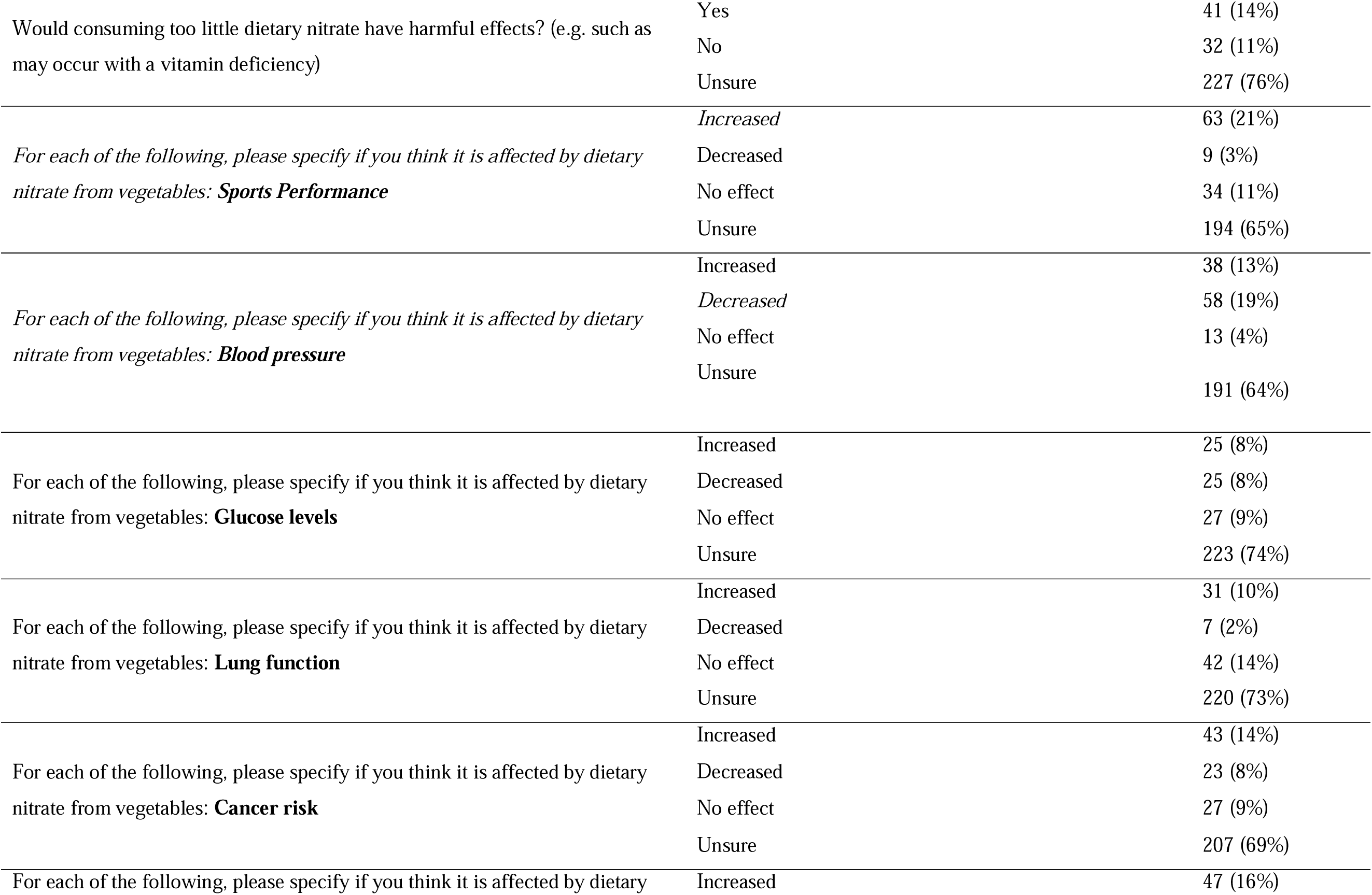
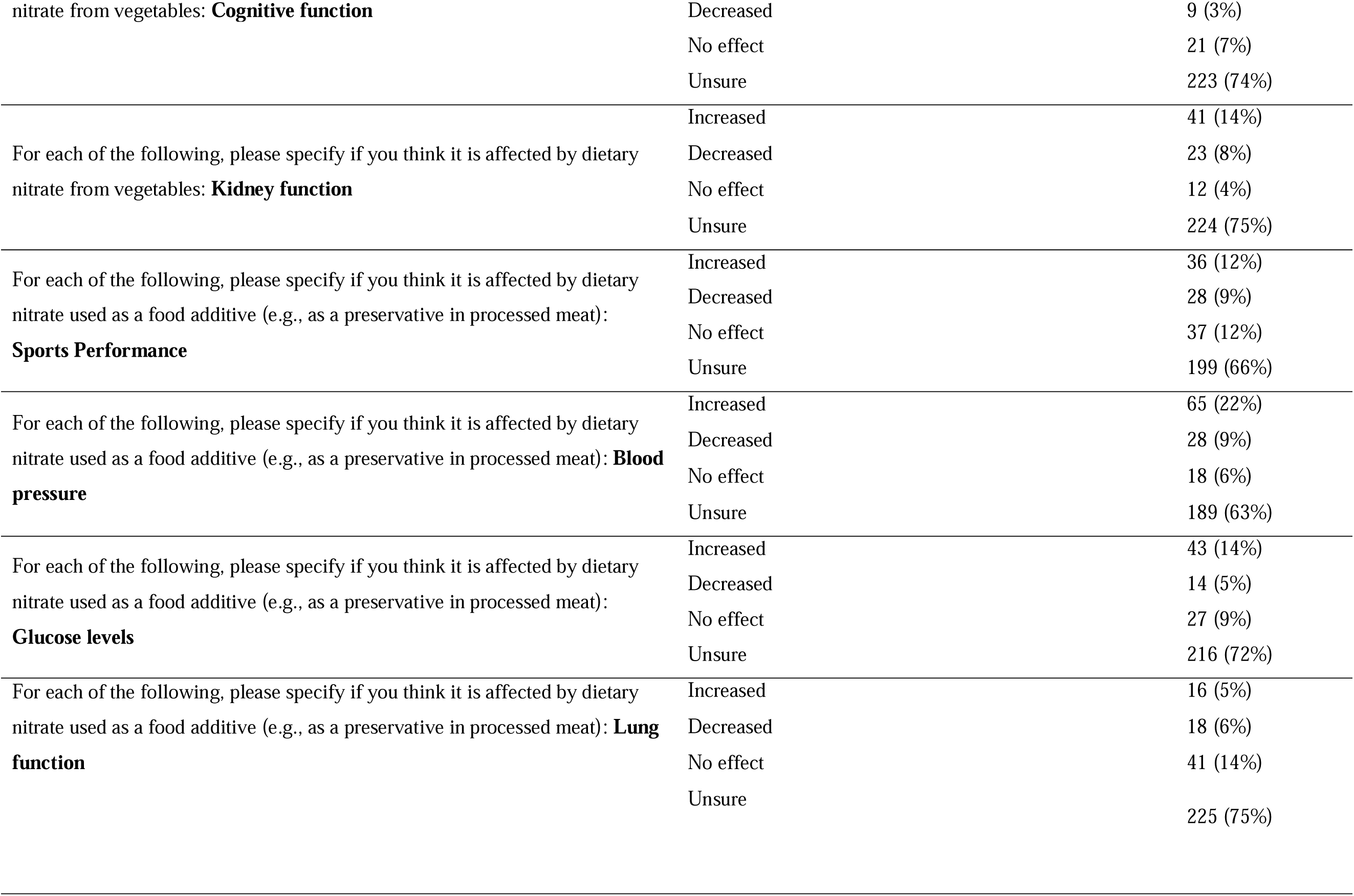
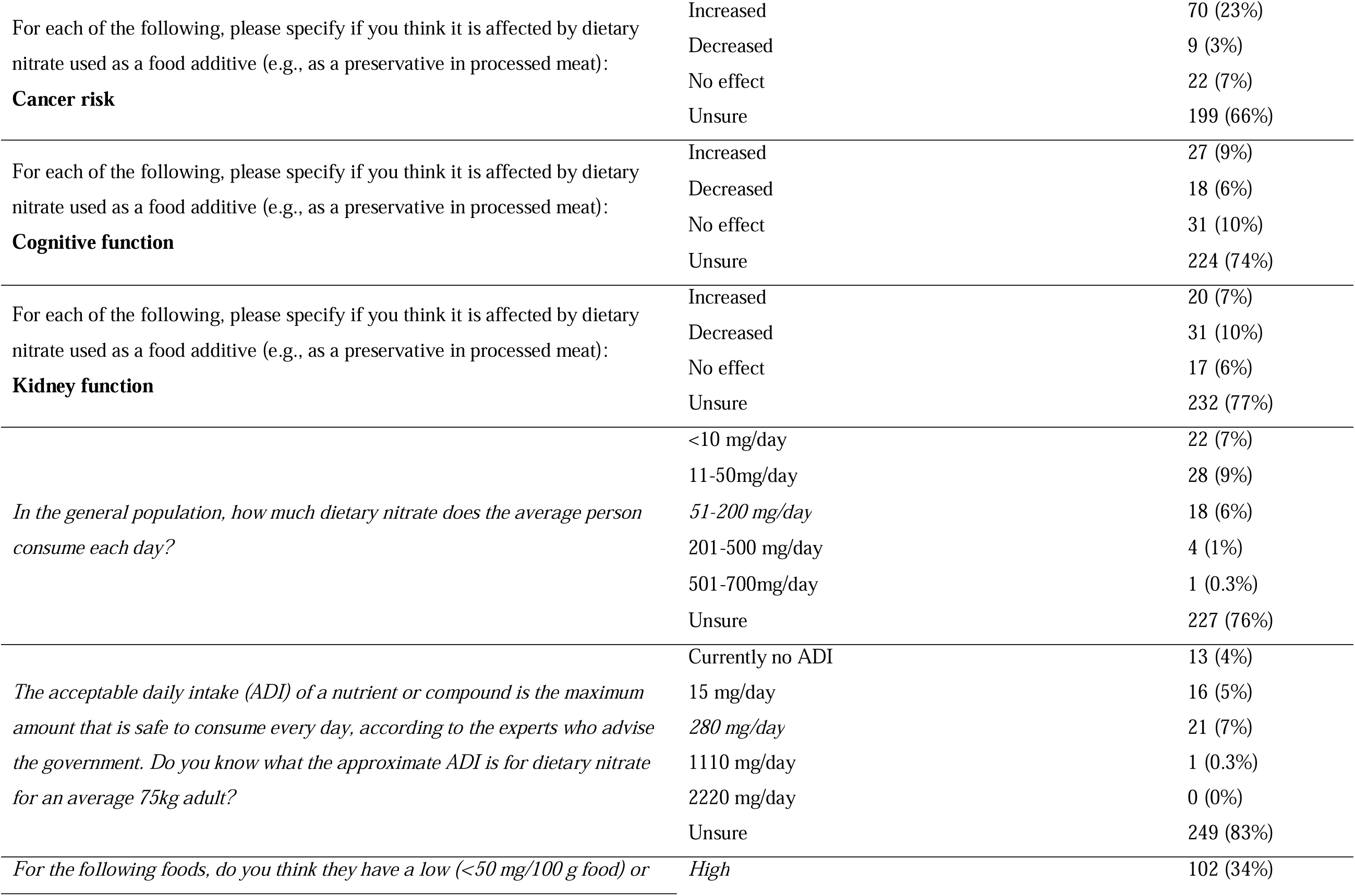
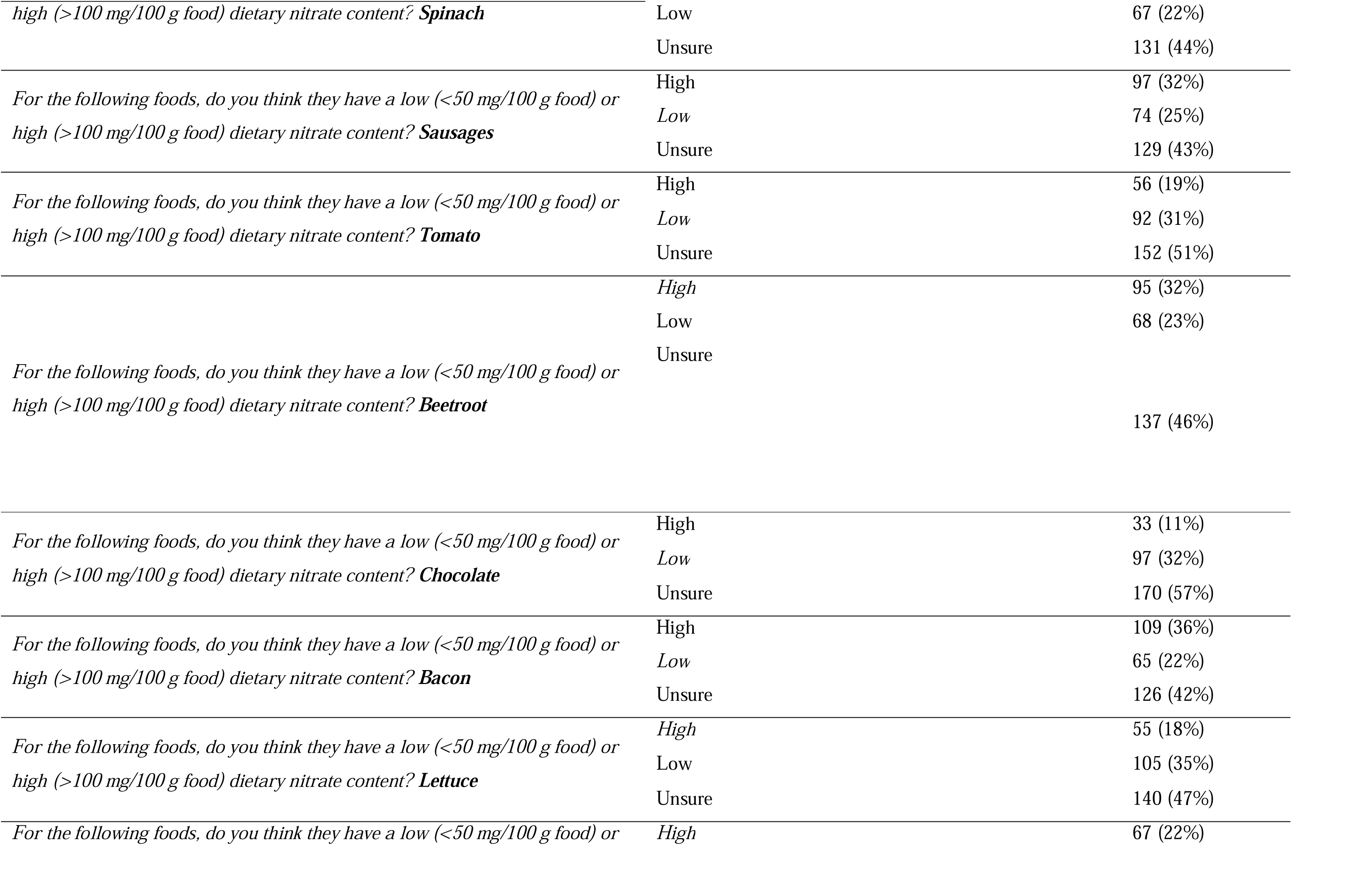
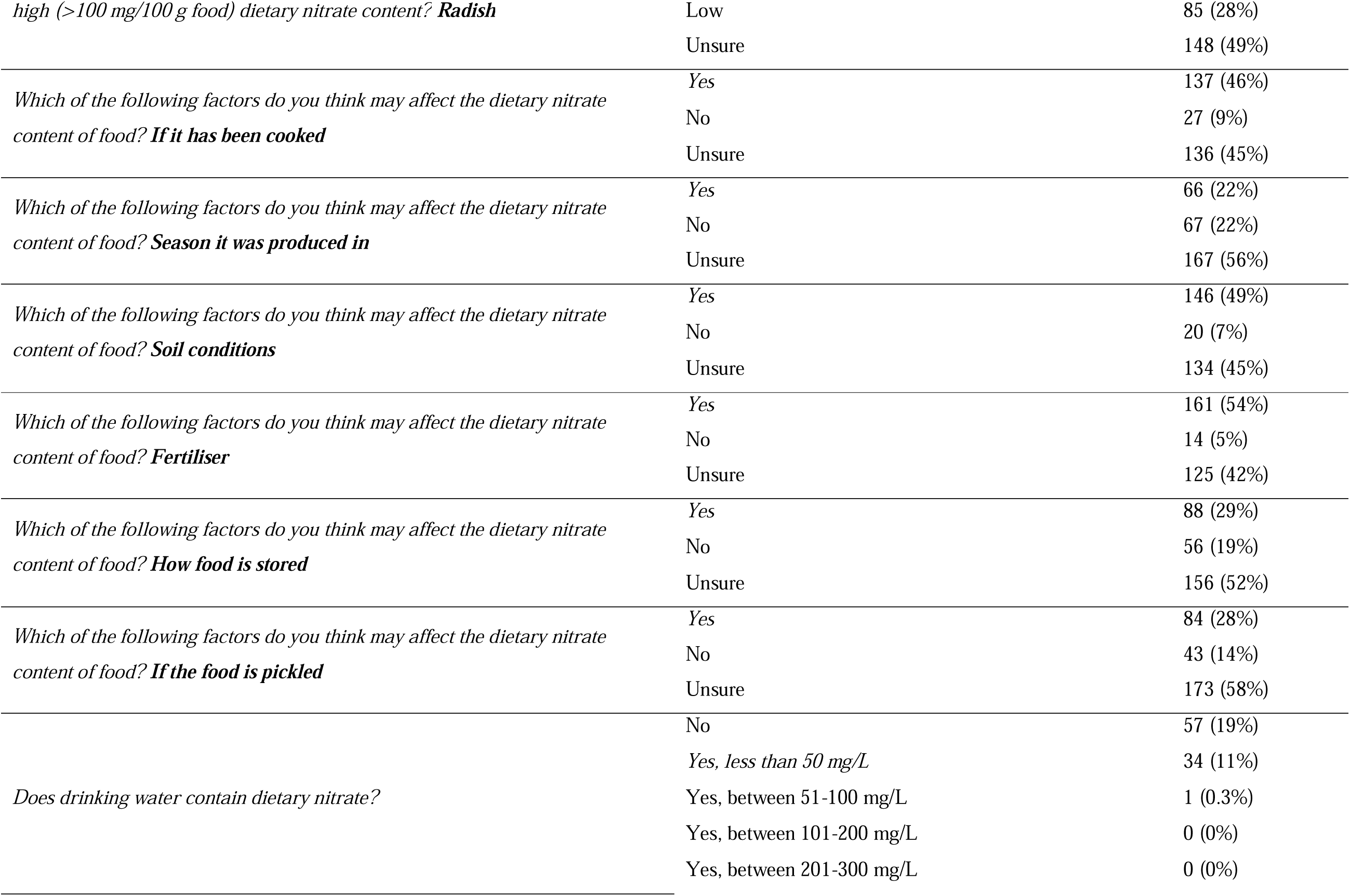
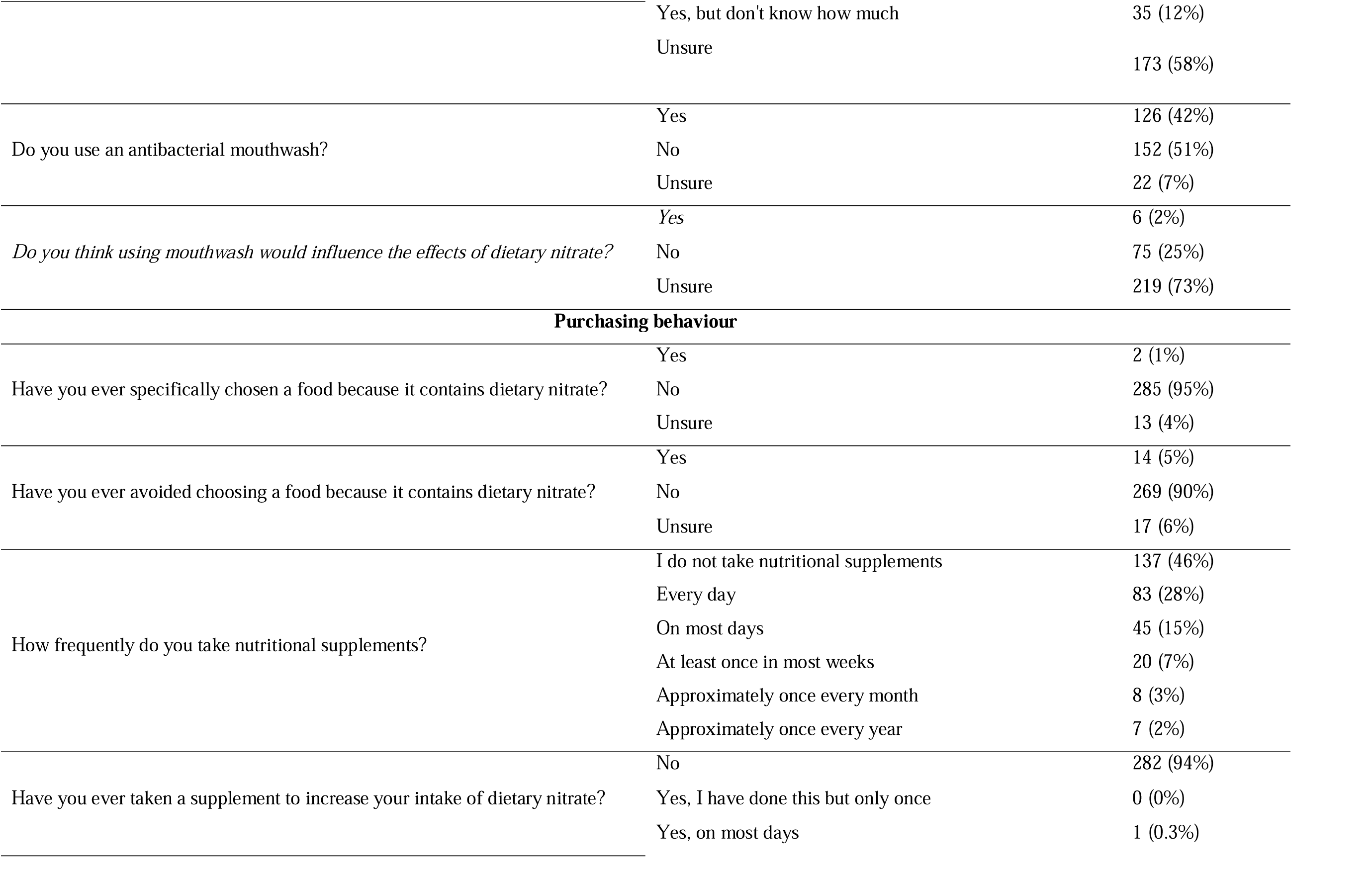
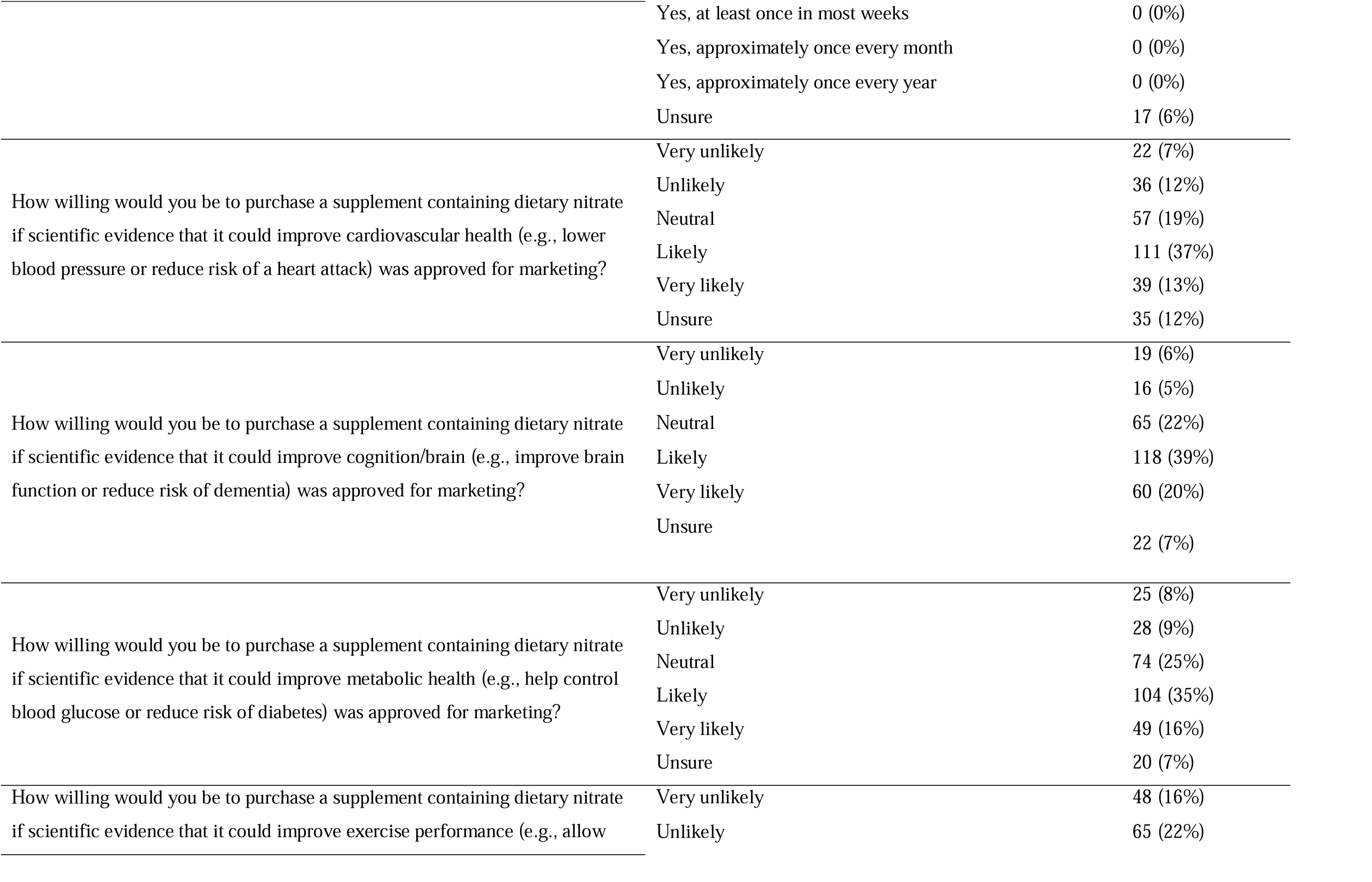
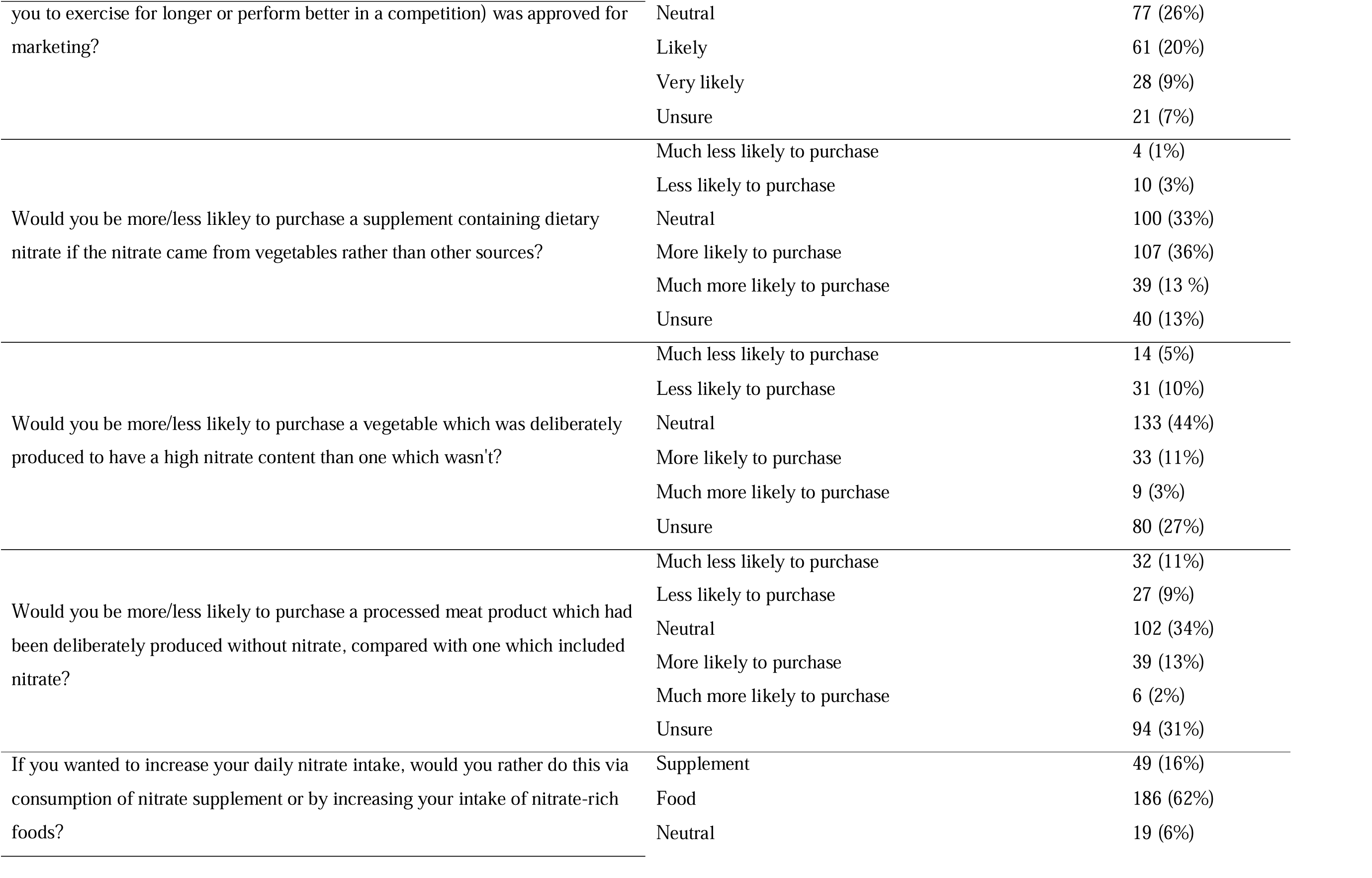
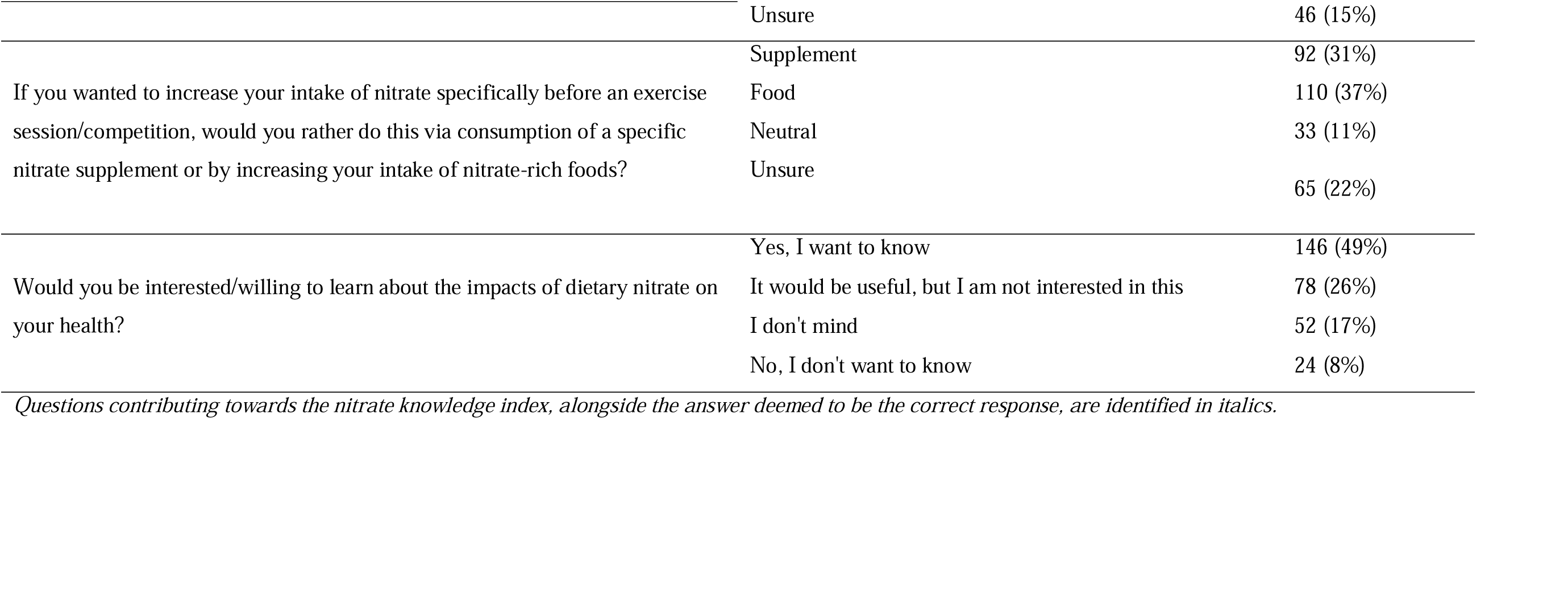
Nitrate knowledge and beliefs in the overall cohort.

### Calculation of Nitrate Knowledge Index

Similar to our previous research within nutrition professionals ^(27)^, a 21-point index was derived to provide a quantitative measure of overall knowledge about dietary nitrate. We identified questions where there was unambiguous evidence for a correct answer. In such cases, participants were awarded one point for correct responses and zero points for incorrect responses (italicised in Table 2). Questions where current evidence is inconclusive/for which no correct response is available were excluded from the Index. Data from recent reviews and an expert consensus statement on nitrate informed decision-making on correct/incorrect responses ^(4,10,23,29,30)^.

### Statistical analysis

Data analyses were conducted using SPSS version 28, whilst figures were created using GraphPad Prism. Differences in knowledge/beliefs about dietary nitrate between different population sub-groups was compared using the chi squared test, whilst the Mann-Whitney U Test was used to compare scores on the Nitrate Knowledge Index between population sub-groups. These population sub-groups were defined by age (younger [<40 years] vs. older [≥ 40 years]), gender (male vs. female), ethnicity (white vs. other), education level (lower [GCSE, A Level, vocational, other] vs. higher [undergraduate degree, Master’s degree or PhD]), employment status (employed and self-employed vs. other), household income (lower [<£35,700] vs. higher [≥£35,700]), BMI (healthy [<25 kg/m^2^] vs. overweight and obese [≥ 25 kg/m^2^]), exercise level (lower [do not exercise] vs. higher [other]), level of nutritional education (lower [no nutritional education, unsure and other] vs. higher [secondary school level of nutritional education and above]), and whether participants had previously heard of nitrate (heard of nitrate vs. have not heard of nitrate). P<0.05 was accepted for statistical significance. Raw data are available in an online repository (https://data.ncl.ac.uk/) and can be accessed by contacting the authors.

## RESULTS

### Participant characteristics

A total of 300 participants completed the questionnaire (Table 1). There was a similar percentage of male (49%) and female (50%) participants within the study, distributed across all geographical areas of the UK. Participants were mostly white (84%), the most common age group was 21-40 years (41%), and the most common BMI range was ‘healthy’ (18.5-24.9) (47%). Most commonly, participants reported exercising 1-2 (34%) or 3-5 (34%) times per week. An undergraduate degree was the most common qualification held (34%). The participants reported mixed levels of nutritional education, with around half reporting no nutritional education (49%) and a third (33%) reporting only basic nutritional education at secondary school.

### Overall knowledge and beliefs about dietary nitrate

An overview of participant responses is provided in Table 2. Overall, only 19% of participants had heard of dietary nitrate prior to completing the questionnaire, compared with 66% who had not heard of nitrate and 15% who were unsure if they had heard of nitrate before.

### Health effects of nitrate

Participants were generally unsure as to whether dietary nitrate from vegetables (74%) or as a food additive (67%) would affect human health. Similarly, most of the participants were unsure as to whether dietary nitrate from vegetables or used as a food additive would affect sports performance (vegetables: 65% unsure, food additive: 66% unsure), blood pressure (vegetables: 64% unsure, food additive: 63% unsure), glucose levels (vegetables: 74% unsure, food additive: 72% unsure), lung function (vegetables: 73% unsure, food additive: 75% unsure), cancer risk (vegetables: 69% unsure, food additive: 66% unsure), cognitive function (vegetables: 74% unsure, food additive: 74% unsure) and kidney function (vegetables: 75% unsure, food additive: 77% unsure).

### Nitrate sources and ADI

Knowledge around dietary nitrate intake was typically poor. Most participants (76%) were unsure of the average population intake of dietary nitrate and the nitrate acceptable daily intake (ADI) (83%). Knowledge of dietary sources of nitrate was generally poor, with ∼20-30% of participants correctly identifying spinach, beetroot, lettuce and radish as high in nitrate, and ∼20-30% of participants correctly identifying sausage, tomato, chocolate and bacon as low in nitrate. Around half of the participants were aware that the dietary nitrate content of food is influenced by cooking (46%), soil conditions (49%) and fertiliser (54%) but a smaller percentage were aware of the influence of season it was produced in (22%), how it was stored (29%) and if the food was pickled (28%). Participants were mostly unsure as to whether drinking water contains dietary nitrate (58%) and whether use of mouthwash would influence the effects of dietary nitrate (73%).

### Purchasing behaviour

Most participants had not chosen (95%) or avoided (90%) purchasing foods because they contained dietary nitrate. Likewise, most had not consumed supplements to increase their intake of dietary nitrate (94%). Around half of all participants reported that they would be likely or very likely to purchase a supplement containing dietary nitrate if scientific evidence demonstrated it could improve cardiovascular health (50%), cognition (59%) and metabolic health (51%), but a smaller percentage were likely or very likely to purchase a nitrate-containing supplement to improve exercise performance (30%). Participants reported that they were more likely or much more likely to purchase a supplement containing dietary nitrate if it came from a vegetable rather than other sources (49%), whereas they were generally neutral with regards to whether they would purchase a vegetable (44%) or processed meat products (34%) which were deliberately produced to have a high dietary nitrate content. Interestingly, most participants reported that they would rather increase their intake of dietary nitrate via consumption of nitrate-rich foods (62%) rather than supplements (16%). Responses were more varied in relation to consuming nitrate prior to exercise, with 37% and 31% of participants preferring to increase dietary nitrate intake via food and supplements, respectively. There was a willingness to learn about the impact of dietary nitrate on health, with 49% of participants stating they would like to know more.

### Differences in knowledge and beliefs in different population groups

There were some significant differences in knowledge and beliefs about dietary nitrate between different participant sub-groups, which are highlighted below and in **Supplementary Table 1**.

### Age

Knowledge/beliefs around the health effects of dietary nitrate consumption as a food additive differed according to age. Specifically, participants aged ≥ 40 years were more likely to believe that nitrate provided as a food additive is harmful compared with those < 40 years (25% vs. 9%; *p*=0.002). Those aged ≥ 40 years were also more likely to perceive *excess* consumption of dietary nitrate as a food additive as harmful, compared with those < 40 years (39% vs. 21%; *p*=0.004). Beliefs around the physiological effects of dietary nitrate consumption were generally consistent between older and younger adults, although participants aged ≥ 40 years were more likely to believe that nitrate intake from vegetables increased cancer risk (≥ 40 years: 17%, < 40 years: 11%, p=0.006). A larger proportion of those aged ≥40 years believed that the way in which food was stored could influence its nitrate content compared with those aged < 40 years (33% vs 25%; *p*=0.02).

With regards to purchasing behaviour, a greater percentage of participants aged ≥ 40 years compared with < 40 years had not specifically chosen a food to increase dietary nitrate intake (98% vs. 91%; *p*=0.008). Similarly, those aged ≥ 40 years were marginally more likely to have avoided a specific food because it contains dietary nitrate than those aged < 40 years (7% vs. 2%; *p*=0.019). A greater percentage of participants aged ≥ 40 years had not taken a supplement to increase dietary nitrate compared with those aged < 40 years (97% vs. 90%; *p*=0.03). Interestingly, participants aged < 40 years were more likely to state that they would rather increase their dietary nitrate intake prior to exercise/competition via a supplement (43% vs. 22%; *p* < 0.001), whereas participants aged ≥ 40 years had a greater preference to consume whole foods to increase their nitrate intake prior to exercise compared with those < 40 years (44% vs. 26%).

### Gender

A greater proportion of male compared with female participants believed that the effects of dietary nitrate on health differ depending on the type of food it is in (7% vs 1%; *p*=0.012). Beliefs around the physiological effects of dietary nitrate consumption were generally consistent between those of different genders, although males were more likely than females to believe that dietary nitrate consumption as a food additive increased exercise performance (17% vs. 7%; *p*=0.038). A greater proportion of male participants believed that the use of antibacterial mouthwash would influence the effects of dietary nitrate compared with female participants (4% vs 0%; *p*=0.041). Male participants were more likely or much more likely than female participants to purchase a nutritional supplement containing dietary nitrate to improve exercise performance (36% vs. 24%; *p* < 0.001) and were also more likely or much more likely to purchase a vegetable which was deliberately produced to have a high nitrate content (19% vs. 9%; *p*=0.024).

### Ethnicity

Beliefs around the physiological effects of dietary nitrate from vegetables were generally consistent between those of different ethnicities. However, a greater percentage of participants from ethnic minority groups believed that dietary nitrate from vegetables either had either no effect on cancer risk (19% vs. 7%) or decreased risk (13% vs. 7%; *p*=0.016) compared with those of white ethnicity. A larger percentage of participants from ethnic minority groups compared with white ethnicity perceived dietary nitrate as a food additive to decrease sports performance (21% vs. 7%; *p*=0.016), increase blood pressure (36% vs. 19%; *p*=0.051) and increase glucose levels (28% vs. 12%; *p*=0.043). Participants from ethnic minority groups also showed some indications of better knowledge about nitrate on specific questions, compared with white participants. For example, a greater percentage of participants from ethnic minority groups minority compared with white participants were aware that lettuce is high in nitrate (32% vs. 16%; *p*=0.008), and correctly identified the nitrate content of drinking water (26% vs. 9%; *p*=0.009). Participants from ethnic minority groups were also more likely than those of white ethnicity to want to learn more about the impact of dietary nitrate on health (68% vs. 45%; *p*=0.037).

### Education

A greater proportion of participants with higher compared with lower-level educational qualifications believed that the effects of dietary nitrate on health did not differ depending on the type of food it is in (4% vs 0%; *p*=0.039). Beliefs around the physiological effects of dietary nitrate consumption were generally consistent between those with different qualifications, although participants with a higher-level qualification were more likely than those with a lower level qualification to believe that dietary nitrate consumption via a food additive increased cognitive function (12% vs. 6%; *p*=0.036).

### Employment status

There was a greater percentage of employed compared with non-employed participants who believed that spinach (41% vs. 23%; *p*=0.006), tomato (24% vs 10%; *p*=0.01), beetroot (38% vs. 21%; *p*=0.008) and radish (28% vs. 14%; *p*=0.015) are high in nitrate, and that bacon is low in nitrate (27% vs 13%; *p*=0.01). A greater percentage of employed compared with non-employed participants believed that antibacterial mouthwash does not impact the effects of dietary nitrate in the body (31% vs. 15%; *p*=0.008). A greater percentage of employed compared with non-employed participants stated that they would rather use a dietary supplement to increase their nitrate intake prior to exercise (34% vs. 25%; *p*=0.034).

### Income

Participants with higher compared with lower income were more likely to want to use a dietary supplement to increase their nitrate intake prior to exercise (36% vs. 25%), whereas those with a lower income were more likely to want to consume food to increase dietary nitrate prior to exercise/competition (46% vs. 28%; *p*=0.002).

### BMI

Beliefs around the physiological effects of dietary nitrate consumption were generally consistent between those with different BMIs, although those with a lower compared with higher BMI were more likely to believe that dietary nitrate intake from vegetables (18% vs. 6%; *p*=0.008) and as a food additive (28% vs. 14%; *p*=0.031) increases blood pressure.

### Exercise level

A greater percentage of those engaging in exercise compared with no exercise perceived spinach to be high in nitrate (38% vs. 25%; *p*=0.01). A greater percentage of participants not engaging compared with engaging in exercise perceived no effect of cooking (17% vs. 6%; *p*=0.005) or season (29% vs. 19%; *p*=0.046) on the nitrate content of food. Those who engage in exercise were more likely to want to learn more about dietary nitrate than those that do not (54% vs 36%; *p*=0.035).

### Nutritional education

Those with a higher compared with lower level of nutritional education were more likely to have heard of dietary nitrate (27% vs. 13%; *p*=0.002). A greater percentage of participants with a higher compared with lower level of nutritional education believed that dietary nitrate from vegetables increases sports performance (28% vs. 16%; *p*=0.047) and cognitive function (23% vs 10%; *p*=0.02), and that dietary nitrate as a food additive increases cognitive function (14% vs. 5%; *p*=0.018). A greater percentage of participants with a higher compared with lower level of nutritional education perceive soil content to influence the nitrate content of foods (53% vs. 45%; *p*=0.007).

### Prior knowledge of nitrate

Knowledge/awareness of nitrate prior to undertaking this survey had the biggest impact on responses to the questionnaire. Participants who had heard of nitrate prior to undertaking this survey were more likely (than those who had not heard of nitrate) to believe that the health effects of nitrate differ depending upon the type of food it is in (14% vs. 2%; *p*<0.001), believing that nitrate from vegetables is beneficial for health (45% vs. 14%; *p*<0.001) but that consumption of nitrate as a food additive is harmful for health (40% vs. 10%; *p*<0.001) (for beliefs about on individual health outcomes, see **Supplementary Table 1**). Participants who had heard of nitrate were generally (although not always) better at identifying foods with higher and lower nitrate content. These individuals were also more likely to believe that the nitrate content of food could be influenced by whether it had been cooked (55% vs. 45%; *p*=0.031), the soil conditions (71% vs. 42%; *p*<0.001), use of fertiliser (74% vs. 51%; *p*=0.001), how it is stored (41% vs. 26%; *p*=0.008) and if the food is pickled (40% vs. 24%; *p*=0.019).

Participants who had heard of nitrate were more likely to correctly identify that drinking water typically contains less than 50 mg/L nitrate (26% vs. 8%; *p*<0.001). Participants who had heard of nitrate more commonly stated that they were ‘very likely’ to purchase a supplement containing nitrate if scientific evidence that it could improve cardiovascular health was approved for marketing (21% vs. 11%; *p*=0.042) and that they were ‘much more likely’ to purchase a supplement where the nitrate came from vegetable sources (26% vs. 9%; *p*=0.007). Similarly, participants who had heard of nitrate more commonly stated that they were ‘less likely’ to purchase a vegetable deliberately produced to have a high nitrate content (17% vs. 8%; *p*=0.024) and ‘more likely’ to purchase a meat product produced without nitrate (29% vs. 9%; *p*<0.001) than those who had not heard of nitrate. Sub-group analyses (analyses by participant characteristics) were run separately in participants who had/had not heard of nitrate as exploratory analyses, and are presented in Supplementary Table 1.

### Nitrate Knowledge Index

Overall knowledge about dietary nitrate was quantified using a Nitrate Knowledge Index. In the entire cohort, the median (IQR) score for the Nitrate Knowledge Index was 5 (8). Knowledge Index stratified by participants demographics is presented in Figure 1. There were no significant associations between age (*p*=0.558), gender (*p*=0.558), ethnicity (*p*=0.52), level of qualification (*p*=0.978), income (*p*=0.535), BMI (*p*=0.246) or exercise (*p*=0.377) and the Nitrate Knowledge Index. However, there was a significant association between employment status and the Nitrate Knowledge Index, with those in employment achieving a higher score compared to those not in employment (Nitrate Knowledge Index (median (IQR)): 5 (7) vs. 4 (7); *p*=0.0007). Nutritional education was also significantly associated with the Nitrate Knowledge Index score, with those with a higher nutritional education achieving a higher score than those with a lower nutritional education (Nitrate Knowledge Index (median (IQR)): 6 (7) vs. 4 (8); *p*=0.012). In addition, participants who had previously heard of nitrate before completing the questionnaire had significantly higher Nitrate Knowledge Index scores than those who had not previously heard of nitrate (median (IQR): 9 (8) vs. 4 (7); *p*<0.001).

**Figure 1.**
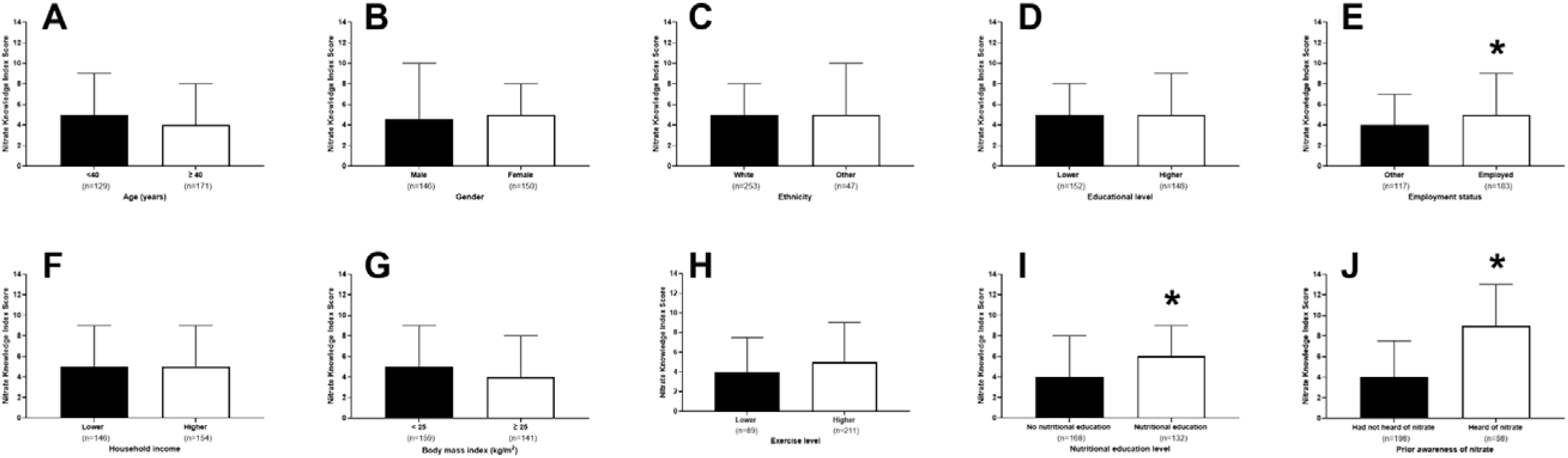
Nitrate Knowledge Index scores in different sociodemographic groups. Analyses were stratified by age (panel A; younger [<40 years] vs. older [≥ 40 years]), gender (panel B; male vs. female), ethnicity (panel C; white vs. other), education level (panel D; lower [GCSE, A Level, vocational, other] vs. higher [undergraduate degree, Master’s degree or PhD]), employment status (panel E; employed and self-employed vs. other), household income (panel F; lower [<£35,700] vs. higher [≥£35,700]), BMI (panel G; healthy [<25 kg/m^2^] vs. overweight and obese [≥ 25 kg/m^2^]), exercise level (panel H; lower [do not exercise] vs. higher [other]), level of nutritional education (panel I; lower [no nutritional education, unsure and other] vs. higher [secondary school level of nutritional education and above]), and prior knowledge of nitrate (panel J; had not heard of nitrate vs. heard of nitrate). Data presented are median (IQR). * = significant difference (p<0.05) between groups. Individuals who were employed, with higher nutritional education, and who had heard of nitrate prior to completing the questionnaire showed significantly greater knowledge of dietary nitrate.

## DISCUSSION

In this study, we set out to evaluate knowledge and beliefs about dietary inorganic nitrate in a representative sample of adults from the United Kingdom. We found that knowledge of dietary nitrate was generally poor, with only one fifth of the participants having heard of this compound prior to completing the questionnaire. In comparison, in a previous study from our group evaluating knowledge and beliefs about dietary nitrate amongst 125 UK-based nutrition professionals, >70% of participants had previously heard of dietary nitrate ^(27)^. We evaluated overall knowledge about dietary nitrate via a 21-point Nitrate Knowledge Index. Median score for the Knowledge Index was 5 in the entire cohort, reflecting typically poor knowledge about this compound. In comparison, in our previous study in nutrition professionals in which nitrate knowledge was evaluated via a 23-point Index (the higher total score available was due to the addition of two questions related to metabolism of nitrate, which was not considered to be relevant in this investigation), the group median score was 12.

In the current study, a minority of participants were able to correctly identify health effects associated with nitrate. Indeed, there appeared to be a general inability to distinguish between scientifically proven effects of nitrate (e.g., blood pressure reduction ^(10,14,31)^ or improved exercise performance ^(23–25)^) and other physiological effects with limited/no supporting evidence (e.g., lung or kidney function). This suggests poor dissemination of current nitrate-related knowledge outside of scientific communities. Only around 1/3 of participants were able to correctly identify foods with a higher or lower dietary nitrate content and therefore would struggle to make informed decisions about the purchase of foods containing this compound. A larger percentage of participants (∼50%) were able to correctly identify some factors impacting the nitrate content of food (e.g., cooking, soil conditions, and use of fertiliser). However, this could reflect general knowledge of factors the impact the nutritional properties of food, rather than knowledge specific to nitrate. In contrast, in our previous study in nutrition professionals, knowledge about the physiological effects of nitrate, food sources of this compound, and factors affecting its content was much greater (typically ∼50% of nutrition professionals gave correct responses for questions on these factors).

Previous research suggests that overall nutritional knowledge in both students ^(32)^ and adults ^(33,34)^ in the UK population is typically low, which mirrors the findings seen here for a specific nutritional compound, dietary nitrate. There is some previous evidence to suggest variations in overall nutritional knowledge between different sociodemographic groups, with men typically having lower nutritional knowledge than women, and individuals with lower education levels or a lower socioeconomic status also possessing lower levels of nutritional knowledge ^(33)^. Although we found some variation in response to individual questions within these groups, we did not find any overall difference in Nitrate Knowledge Index scores by gender, overall education levels, or household income (our closest proxy for socioeconomic status). However, we found that individuals who reported some previous nutritional education scored on average 2 points higher on the Nitrate Knowledge Index than those without any previous nutritional education. This is broadly consistent with the findings from our previous investigation in which we found that individuals with higher level nutritional qualifications (possession of a Master’s/PhD) typically had greater knowledge of nitrate than individuals with lower level qualifications (undergraduate degree) ^(27)^. Interestingly, in the current study, individuals who were employed or self-employed also had significantly greater (median 1 point higher) scores on the Nitrate Knowledge Index than individuals who were retired, unemployed or students. This agrees with some previous research in Australian adults, which showed greater overall nutritional knowledge in employed versus unemployed adults ^(35)^. It is possible that employed individuals may have had greater nutritional education or exposure to relevant information as part of their employment. The greatest differences in Nitrate Knowledge Index scores were observed between those had vs. had not heard of dietary nitrate. Specifically, those who had heard of nitrate scored on average 5 points higher on the Nitrate Knowledge Index score than those who had not heard of dietary nitrate. Differences in knowledge were particularly apparent when identifying the health effects of dietary nitrate, the dietary nitrate content of different foods and factors which influence the dietary nitrate content of foods.

Our study provides new information on whether an individual’s knowledge or beliefs about dietary nitrate impacts, or could impact, their behaviour when purchasing foods. Almost all participants reported that they had not deliberately purchased, or avoided purchasing, a food because of its nitrate content. This suggests that simply marketing a food as higher/lower nitrate (which is sometimes the case for processed meat products which have been deliberately prepared without nitrate due to perceived health risks of this compound) may not impact consumer behaviour. Similarly, most participants had not previously purchased a nitrate-rich supplement to increase their intake of this compound, although around half of the participants suggested they would be likely or very likely to do this if there was evidence to suggest this could have cardiovascular, metabolic or cognitive benefits. Therefore, if such claims were approved for use in marketing products, they could have important implications for the sale of such supplements. It is interesting to note that most participants suggested that they would rather increase their intake of nitrate via consumption of nitrate-rich foods rather than supplements. This information could help with the design of interventions and public health campaigns to augment nitrate intake by the public if a sufficient evidence-base was to emerge to support the health benefits of such an approach. Potential advantages of increasing nitrate intake via food versus supplements, previous highlighted by our group ^(3)^, include lower cost, greater variety, and provision of fibre which is often lacking in nitrate-based supplements but can have health benefits ^(36,37)^. Nevertheless, whilst foods are often preferred to supplements (by both individuals and nutritionists/dietitians), there may still be a potential role for supplements under certain circumstances (e.g., athletes looking to consume a high nitrate bolus pre-competition) ^(38)^.

Given the typically poor knowledge of dietary nitrate and associated health benefits, it may be pertinent to develop and/or optimise public health education strategies. This is particularly relevant given the high rates of cardiovascular disease ^(39)^ and dementia ^(40)^ in the UK, and the evidence that dietary nitrate may improve markers of cardiovascular ^(10–14)^ and brain ^(15–18)^, health. Although improved nutritional knowledge does not guarantee behaviour change, it can contribute towards, and facilitate, such changes ^(41)^. Given widespread use of social media websites/applications, such channels may represent an easy-to-use, low-cost, direct way for nutritional educators to reach a relevant audience ^(42)^. Such strategies could be tailored for different sociodemographic groups to maximise impact and knowledge dissemination.

From an international perspective, the findings of limited knowledge about dietary nitrate in the general public may only be relevant to those countries who practice similar legislation to the UK, such as countries within the European Union. The UK has not approved any nitrate-related health claims and, as such, there is limited marketing of this nature. In other regions of the world in which companies are allowed to advertise the health effects of nitrate (e.g. via television and social media), knowledge about dietary nitrate could be greater and future research is required to measure this.

### Strengths and limitations

This study includes a relatively large sample, similar to/greater than the number of participants in previous investigations into nutritional knowledge ^(27,35,43,44)^. The dataset produced could serve as a reference point for future investigations into nitrate knowledge in different populations or to evaluate change in knowledge in the public over time. We matched our participant characteristics to the wider UK population by age, gender and ethnicity. However, we were unable to ensure all characteristics of our participants were representative of the UK population, including education and employment status, both of which can influence nutritional knowledge/habits ^(35)^. Considering this was a nutrition-based questionnaire, it is possible that respondents may have had a greater interest in nutrition compared with non-respondents. However, as the key finding ultimately reflects a limited knowledge of nitrate, the prior interests of participants are unlikely to have significantly affected the general ‘take home’ message from this study. Another limitation of the study is that few participants (n=58) had heard of dietary nitrate prior to completing the questionnaire. It is therefore possible that some responses from individuals who had not heard of nitrate were educated guesses based on general nutritional knowledge/beliefs. Additional analysis demonstrated that those that had heard of nitrate performed significantly better than those that had not heard of nitrate. A final strength of this questionnaire is that it was underwent considerable pilot testing with members of the public, which maximised comprehensibility^(45)^.

### Conclusion

This study provides new insight into knowledge and beliefs about dietary nitrate in the general population of the United Kingdom. Overall, results show that knowledge about dietary nitrate is poor, and notably lower than previously observed for nutrition professionals. Greater education around this compound may be valuable to help individuals make more informed decisions about their consumption of nitrate-containing foods. This could occur as part of broader efforts to increase nutritional knowledge in the population, which could be an important strategy to mitigate risk of diet-associated diseases including cardiovascular disease, diabetes, cancer and dementia ^(46–48)^.

## Data Availability

Raw data are available in an online repository (https://data.ncl.ac.uk/) and can be accessed by contacting the authors.

## Notes

### Competing Interest Statement

The authors have declared no competing interest.

### Funding Statement

This research received no specific grant from any funding agency, commercial or not-for-profit sectors.

### Author Declarations

This study was approved by the Newcastle University Ethics Committee (REF: 26332/2022).

